# Survey of the Landscape of Society Practice Guidelines for Genetic Testing of Neurodevelopmental Disorders

**DOI:** 10.1101/2024.02.22.24302957

**Authors:** Siddharth Srivastava, Jordan J. Cole, Julie S. Cohen, Maya Chopra, Hadley Stevens Smith, Matthew A. Deardorff, Ernest Pedapati, Brian Corner, Julia S. Anixt, Shafali Jeste, Mustafa Sahin, Christina A. Gurnett, Colleen A. Campbell, the Intellectual and Developmental Disabilities Research Center (IDDRC) Workgroup on Advocating for Access to Genomic Testing

## Abstract

Genetic testing of patients with neurodevelopmental disabilities (NDDs) is critical for diagnosis, medical management, and access to precision therapies. Because genetic testing approaches evolve rapidly, professional society practice guidelines serve an essential role in guiding clinical care; however, several challenges exist regarding the creation and equitable implementation of these guidelines. In this scoping review, we assessed the current state of United States professional societies’ guidelines pertaining to genetic testing for unexplained global developmental delay, intellectual disability, autism spectrum disorder, and cerebral palsy. We describe several identified shortcomings and argue the need for a unified, frequently-updated and easily-accessible cross-specialty society guideline.

## INTRODUCTION

Neurodevelopmental disorders (NDDs) or developmental disabilities are a spectrum of conditions characterized by delay, deviance, and/or dissociation across domains of childhood development (such as motor, problem solving, social-communication, and adaptive skills) ^1^. NDDs are heterogeneous in clinical presentation, resulting in variable definitions. For example, United States law (section 102 of the Developmental Disabilities Assistance and Bill of Rights Act of 2000) defines a developmental disability as a “severe, chronic” condition “attributable to mental or physical impairment,” with onset before the age of 22 years, that is “likely to continue indefinitely,” resulting in “substantial functional limitations in three or more…areas of major life activity,” and requiring lifelong/extended services, supports, or assistance ^2^. In contrast, the Diagnostic and Statistical Manual of Mental Disorders, Fifth Edition (DSM-5) defines NDDs as a group of conditions “characterized by developmental deficits that produce impairments of personal, social, academic, or occupational functioning”, manifesting “early in development”, with a “range of developmental deficits [that] varies from very specific limitations of learning or control of executive functions to global impairments of social skills or intelligence” ^3^. Specific NDDs in the DSM-5 include intellectual disability (ID), global developmental delay (GDD; often a precursor to ID, diagnosed before age 5 years, prior to the age at which one can participate in formal neuropsychological testing), and autism spectrum disorder (ASD). Cerebral palsy (CP), though not explicitly cited by the DSM-5 as an NDD, meets the definition set forth by the Developmental Disabilities Assistance and Bill of Rights Act of 2000.

NDDs are defined by clinical features and behaviors. In the absence of known environmental or acquired causes for NDDs, genetic etiologies are common and include chromosomal copy number variants (CNVs), single-gene disorders, methylation alterations, and trinucleotide repeat expansions ^4^. For varying combinations of GDD/ID/ASD, the prevalence of pathogenic CNVs and monogenic disorders is approximately 10-15% and 30-40%, respectively ^5,6^. For CP, the prevalence of pathogenic CNVs and single-gene disorders is approximately 5% and 20-30%, respectively ^7^.

With the expanding knowledge of genetic causes of NDDs and the rise of next-generation sequencing (NGS) – a technology that rapidly sequences millions of DNA fragments in parallel and uses bioinformatics to analyze the data, allowing for whole genome sequencing (WGS), exome sequencing (ES), and targeted gene panels ^8^ – professional society guidelines pertaining to genetic testing are increasingly important in clinical care. These guidelines, which may also be called practice parameters, are based on comprehensive evidence reviews to inform best practices with respect to diagnosis, management, and treatment ^9^. One of the first guidelines for genetic testing for an NDD originated in 2000, when the American Academy of Neurology (AAN) and Child Neurology Society (CNS) published recommendations for genetic testing for ASD, suggesting karyotype and Fragile X testing ^10^. Since then, other specialty organizations, including pediatrics, genetics, and psychiatry, have formulated their own guidelines for genetic testing of NDDs. Clinicians from these respective specialties who see patients with NDDs often consult society guidelines in deciding whether genetic testing would be helpful, and if so, which genetic test(s) are useful to establish a molecular diagnosis. Payers use society guidelines when writing medical policies to determine insurance coverage for genetic testing for NDDs^11^.

Despite the fact that genetic testing is increasingly important for diagnosing NDDs, a summary of recommendations across professional societies does not yet exist. Therefore, we aimed to fill this gap by conducting a scoping review of the literature on available U.S.-based professional society practice guidelines that contain recommendations for genetic testing of unexplained NDDs. We describe the current practice guideline landscape, highlight challenges, and provide recommendations to improve the efficiency, contemporaneity, cohesiveness, and impact of societal guidelines for genetic testing for NDDs.

For this review, we focused exclusively on GDD/ID, ASD, and CP, as these are the NDDs with primary literature and meta-analyses supporting the diagnostic utility of genetic testing among patients without a clearly identifiable acquired etiology ^6,7,12–14^. We did not include epilepsy, given that it is not conventionally considered an NDD, although this notion is evolving ^15,16^. Individuals who meet criteria for both epilepsy and GDD/ID, ASD, and/or CP are included within the guidelines pertaining to those disorders.

We acknowledge that the spectrum and diagnostic yield of genetic testing for other NDDs, such as attention deficit hyperactivity disorder (ADHD) and specific learning disabilities, are not addressed in this scoping review but should be added in the future as additional literature regarding these disorders is published. We also acknowledge that societal guideline updates may be in progress that are not reflected in this article. The target audience for this review are policy makers and medical societies across multiple medical disciplines, including pediatrics, developmental-behavioral pediatrics, psychiatry, genetics, and neurology.

## METHODS: IDENTIFICATION OF RELEVANT PRACTICE GUIDELINES

We conducted a scoping review including a primary search strategy and manual article selection to identify practice guidelines. In the primary search strategy, we searched articles indexed by PubMed (title, abstract, and keyword search via https://pubmed.ncbi.nlm.nih.gov/) with the following query:

> ("global developmental delay" OR "intellectual disability" OR "autism" OR "cerebral palsy" OR "developmental disabilities") AND ("practice parameter" OR "practice guideline" OR "evidence report" OR "clinical guideline" OR "consensus statement" OR "comprehensive evaluation" or "clinical report" or "practice resource")

Notably, we did not include the term “genetic(s)” in the search query, as society guidelines focusing on multiple aspects of care related to a specific NDD may not include this term(s) in the abstract or as part of MeSH keywords identified by the PubMed search.

We applied inclusion/exclusion criteria to the resulting articles. Inclusion criteria were: statement by a U.S. medical organization/society, focus on an NDD (specifically GDD/ID, ASD, and/or CP), and inclusion of clinical recommendations pertaining to genetic testing. Exclusion criteria were: non-English article; erratum to another article; commentary article; animal, *in vitro*/*in vivo*, biomarker, or other biological study; case report/case series; primary research article; review article or guidelines related to a specific genetic disorder; focus on a study population not of interest; focus on outcomes, management, or diagnostic practices not of interest; lack of clinical recommendations; involvement by an organization outside of the U.S.; retired guideline; guideline replaced by a more contemporary version from the same medical society focusing on the same target population; and guideline/review of guidelines without involvement of a professional organization. The manual article selection phase involved a direct query of all authors to identify any additional articles meeting inclusion/exclusion criteria.

For each article included in the review, we extracted the following data:

- Article metadata
- Society involved and its overall clinical focus
- NDD population of interest
- General focus of the guideline (etiology, diagnosis, management)
- Recommendations pertaining to genetic testing
- Recommendations of ES/WGS as a first-line or second-line test
- Whether the guideline focused on a disorder (such as GDD/ID) or a genetic test (such as ES)

The first author SS manually completed the article search. Both first authors completed the record screening, eligibility review, and data collection. There were no missing data. This study did not require a registered research protocol or statement of approval by an ethical standards committee.

## RESULTS: OVERVIEW OF PROFESSIONAL SOCIETY GUIDELINES

From the primary search strategy, the PubMed query resulted in 531 articles (date of query 2023-08-16). The manual selection process yielded one additional article (AAP 2020 practice parameter on DD ^17^). Of the 532 total articles, nine met inclusion criteria and 523 were excluded. The PRISMA 2020 Flow Diagram is shown in Figure 1.

**Figure 1.**
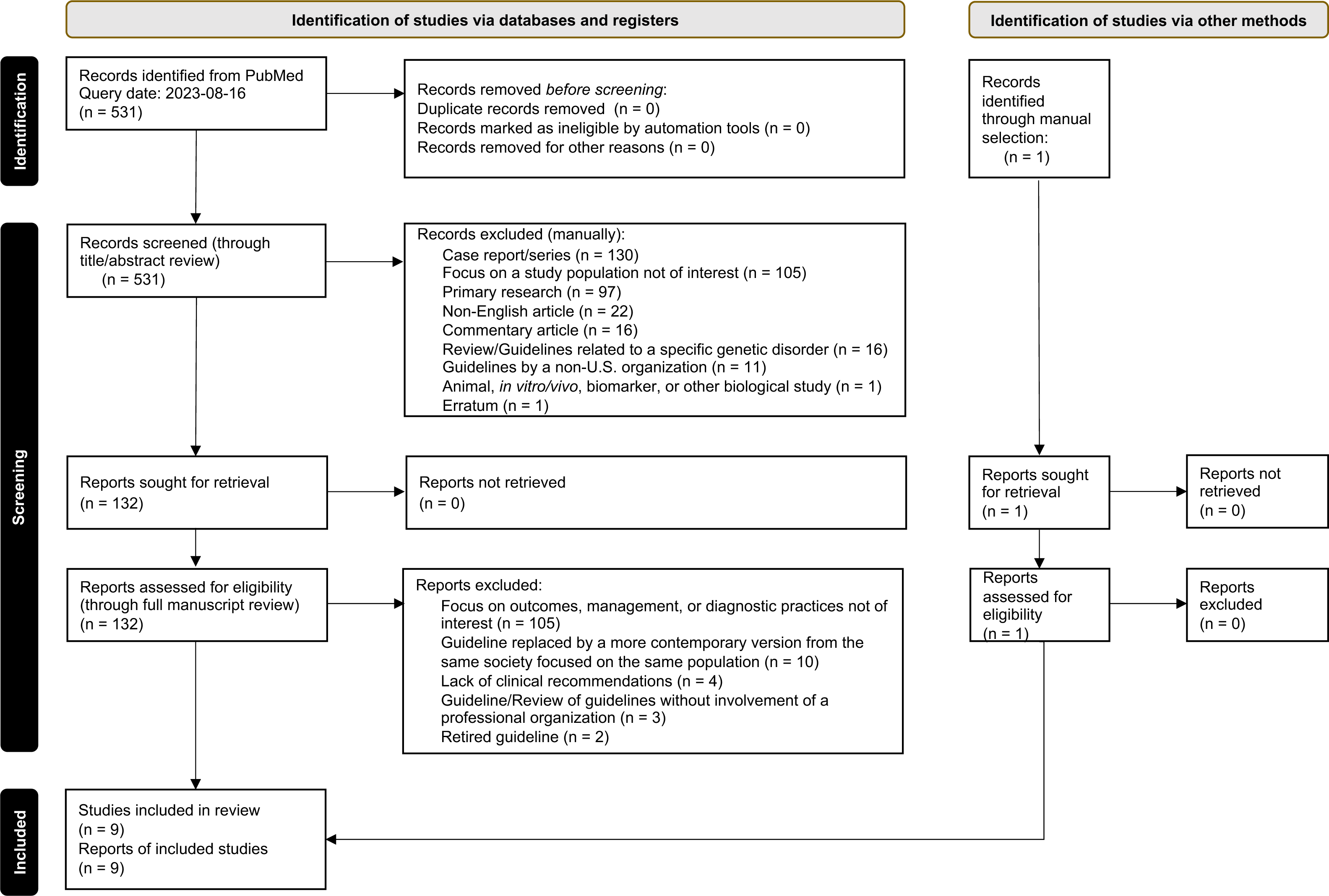
PRISMA diagram showing identification of studies used in the scoping review.

Characteristics of the nine included articles are shown in Table 1 and details of the genetic testing recommendations within each article are outlined in Table 2. The publication years ranged from 2000 to 2022, and only four guidelines were published within the last five years (2018 and onward) ^17–20^. The nine guidelines were from six medical societies representing four specialties (pediatrics, neurology, medical genetics, and psychiatry): American Academy of Child and Adolescence Psychiatry (AACAP), American Academy of Neurology (AAN), American Academy of Pediatrics (AAP), American College of Medical Genetics and Genomics (ACMG), American Society of Human Genetics (ASHG), and Child Neurology Society (CNS). Four guidelines focused exclusively on genetic/etiological evaluation ^5,18,21,22^, while the remaining focused on diagnosis, management, and etiological evaluation. There were four society guidelines relevant to individuals with GDD/ID, published between 2010 to 2021 ^5,17,18,21^. Four society guidelines focused *exclusively* on ASD ^10,20,22,23^, and two guidelines discussed ASD in addition to GDD/ID ^5,17^. With respect to CP, there was one contemporary society guideline, from the American Academy of Pediatrics (AAP) in 2022 ^19^. Out of these nine guidelines, only the ACMG guidelines, published in 2021, recommended WGS or ES as a first- or second-tier test for unexplained NDDs ^18^. We inferred several themes from these guidelines discussed below.

**Table 1.**
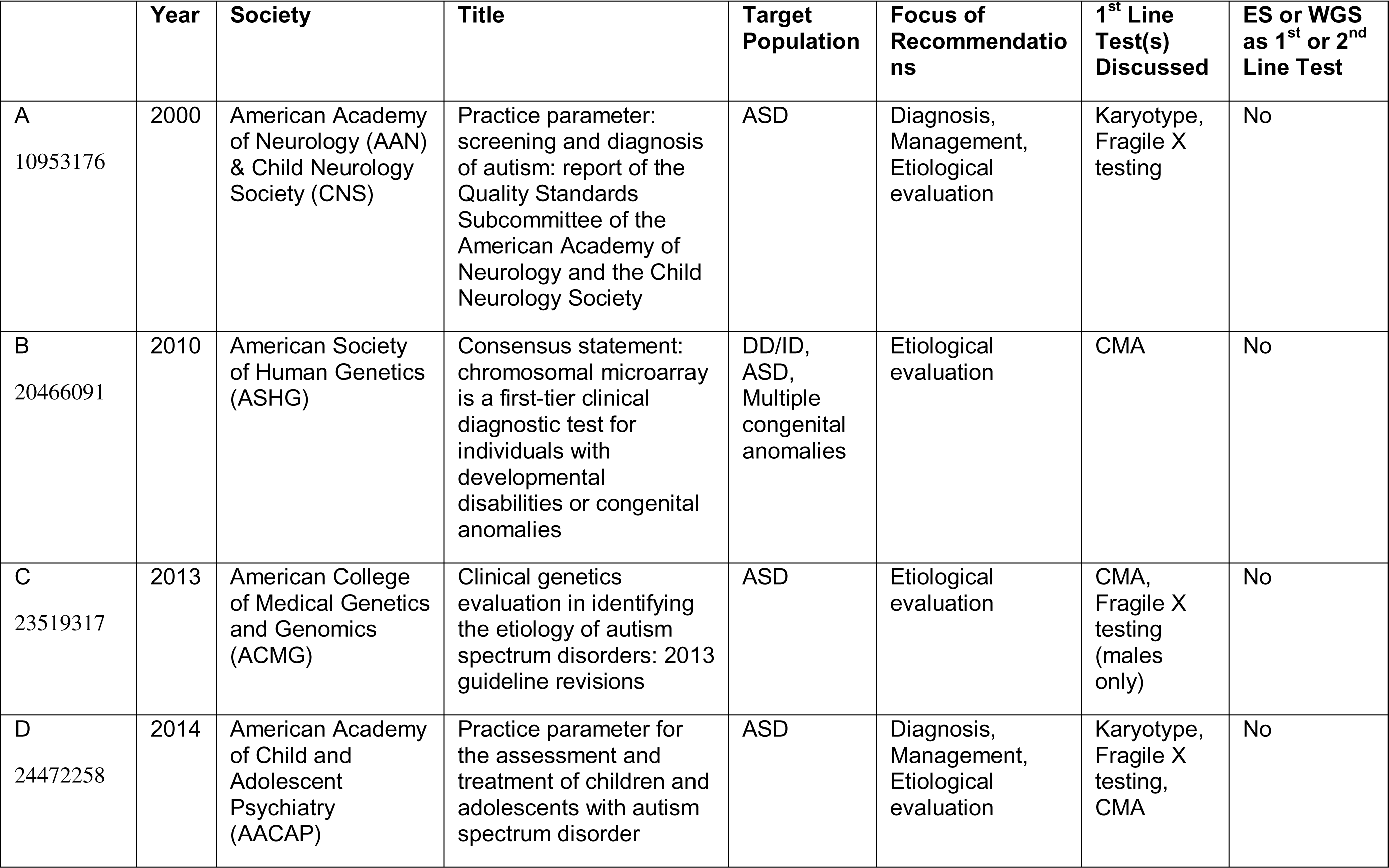
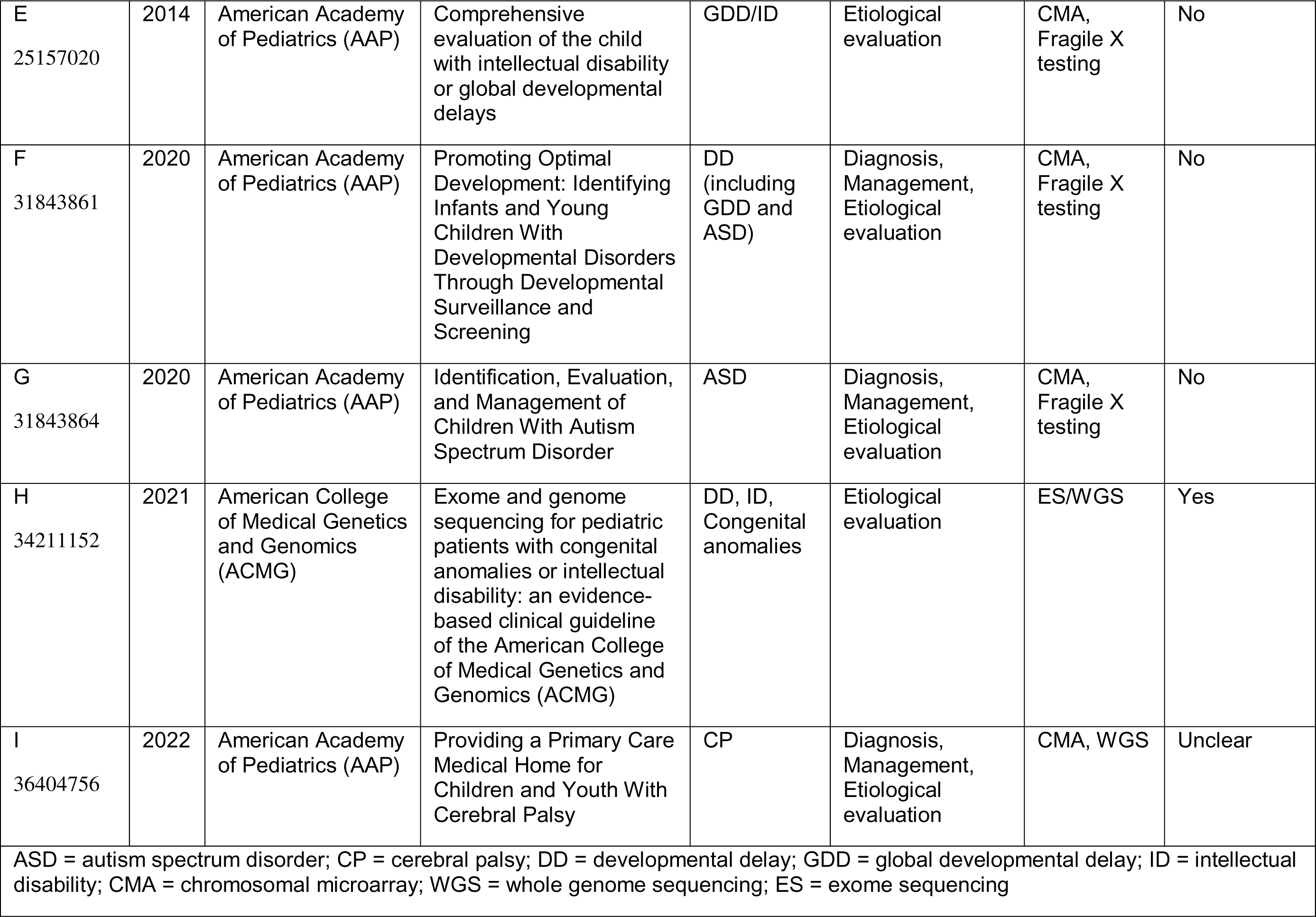
Characteristics of nine professional society guidelines with information on genetic testing for neurodevelopmental disorders. Pubmed ID is included in column on the far left.

**Table 2.** Details of genetic testing recommendations within nine professional society guidelines related to neurodevelopmental disorders. Pubmed ID is included in column on the far left.

|  | Year | Society | Target Population | Detailed Recommendations |
| --- | --- | --- | --- | --- |
| A<br>10953176 | 2000 | American Academy of Neurology (AAN) & Child Neurology Society (CNS) | ASD | 1. Send karyotype and fragile X testing if there is co-occurring ID (or if ID cannot be excluded), family history of fragile X syndrome or undiagnosed ID, or dysmorphic features.<br>2. Send metabolic testing in specific circumstances. |
| B<br>20466091 | 2010 | American Society of Human Genetics (ASHG) | DD/ID, ASD, Multiple congenital anomalies | 1. CMA is a first-tier test for unexplained DD/ID, ASD, or multiple congenital anomalies unless the patient has a recognizable chromosomal syndrome, a family history of a chromosomal rearrangement, or a family history of multiple miscarriages, in which case karyotype is appropriate as a first-tier test.<br>2. If CMA is normal, second-tier testing including fragile X testing, single gene tests, or other molecular test panels should be considered based on clinical presentation. |
| C<br>23519317 | 2013 | American College of Medical Genetics and Genomics (ACMG) | ASD | 1. Perform a three-generation family history with pedigree analysis. Perform comprehensive history and physical evaluation. If a specific syndrome is suspected, send targeted testing. Perform metabolic and/or mitochondrial testing if suggestive clinical indicators.<br>2. If autism is unexplained, send CMA as a first-tier test for all patients and fragile X testing for males.<br>3. Second tier testing as follows: *MECP2* sequencing for all females. *MECP2* duplication testing in males if phenotype is suggestive. *PTEN* testing if head circumference is > 2.5 SD above mean. |
| D<br>24472258 | 2014 | American Academy of Child and Adolescent Psychiatry (AACAP) | ASD | 1. Perform medical assessment, including physical examination, a hearing screen, a Wood's lamp examination, and genetic testing, which may include karyotype, fragile X testing, or CMA. |
| E<br>25157020 | 2014 | American Academy of Pediatrics (AAP) | GDD/ID | 1. Perform comprehensive history and physical examination. If specific diagnosis is certain, inform the family. If specific diagnosis is suspected, send targeted testing.<br>2. If no specific diagnosis is suspected, CMA and fragile X testing should be first-line tests in all. Consider metabolic testing (round 1).<br>3. If family history is suggestive of X-linked disorder, send X-linked ID panel and high-density X-CMA. Consider testing for X-inactivation skewing in the mother of the proband. If the patient is female, send *MECP2* sequencing and deletion/duplication analysis. |
| F<br>31843861 | 2020 | American Academy of Pediatrics (AAP) | DD (including GDD and ASD) | 1. For suspected GDD/ID or ASD, perform CMA and fragile X testing as first-tier tests.<br>2. For suspected GDD/ID, consider metabolic testing if indicated by history and physical examination.<br>3. For suspected GDD/ID, second-tier testing may include ES and gene panels. For suspected ASD, consider genetics consultation. |
| G<br>31843864 | 2020 | American Academy of Pediatrics (AAP) | ASD | 1. Consider immediate referral to clinical genetics to guide genetic evaluation.<br>2. If pursuing first-tier testing oneself, perform comprehensive history and physical examination. If specific diagnosis is suspected, send targeted testing.<br>3. If no specific diagnosis is suspected, discuss and offer CMA and fragile X analysis. If family history is suggestive of X-linked disorder, refer to clinical genetics. If patient is female, consider *MECP2* testing.<br>4. If first-tier tests are unrevealing, consider referral to genetics, workup might include ES. |
| H<br>34211152 | 2021 | American College of Medical Genetics and Genomics (ACMG) | DD, ID, Congenital anomalies | 1. ES/WGS should be a first- or second-tier test in all patients with unexplained DD/ID or congenital anomalies. |
| I<br>36404756 | 2022 | American Academy of Pediatrics (AAP) | CP | 1. Diagnostic evaluation may include CMA or genome sequencing. |

## RESULTS: MANY PROFESSIONAL SOCIETY GUIDELINES ARE OUTDATED

The majority of society-based practice guidelines discussing genetic testing for NDDs (specifically GDD/ID, ASD, and CP) are more than five years old. For example, among the four different society guidelines pertaining to individuals with GDD/ID, the only two published in the past five years were the ACMG 2021 guidelines on developmental delay (DD), ID, and congenital anomalies ^18^ and the AAP 2020 guidelines on DD (including GDD and ASD) ^17^. A 2003 guideline on GDD from the AAN and CNS ^24^ was retired and thus excluded from our scoping review; no updated guideline on GDD from these societies was identified. Among the six guidelines referencing ASD, only two were from the past five years: the AAP 2020 guidelines on DD ^17^ as mentioned above and the AAP 2020 guidelines on ASD ^20^. For CP, there was one recent society guideline, from the AAP in 2022 ^19^. A 2004 guideline on CP from the AAN ^25^ was retired and thus excluded from our scoping review; no updated guideline on CP from the AAN was identified. In summary, out of a total of nine practice guidelines published between 2000 and 2022, only 4/9 (44%) were from the past five years.

## RESULTS: RECOMMENDATIONS DO NOT REFLECT CONTEMPORARY KNOWLEDGE

The most recently published guidelines for an NDD often do not reflect contemporary knowledge and/or availability of modern genetic testing options. For example, ES became available as a clinical test in 2011, thus guidelines published before or around this time (two of the nine guidelines identified) do not reflect the paradigm shift in genomic evaluation (Figure 2). More recently, converging data from different disorders (NDDs or otherwise) supports first-line broad testing with ES or WGS especially to aid in narrowing a differential diagnosis or making a molecular diagnosis. The ACMG 2021 guidelines reflect this through their recommendation of ES/WGS as a first- or second-tier test for patients with DD, ID, or congenital anomalies ^18^.

**Figure 2.**
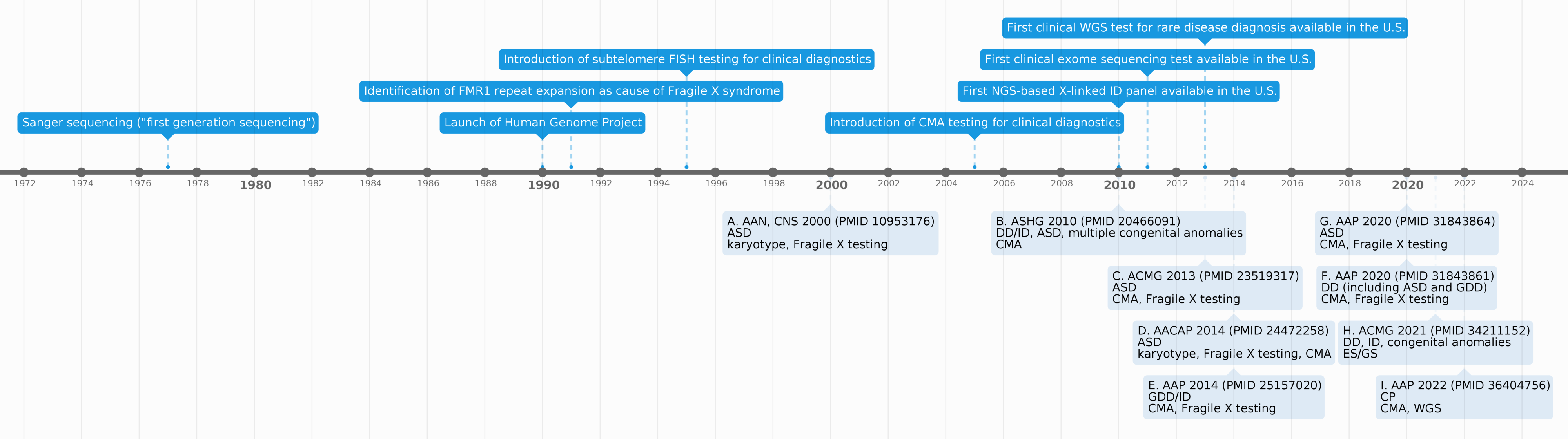
Timeline depicting the advent of different genetic sequencing technologies (top) and year of publication of different NDD practice parameters (bottom). The letter in front of each guideline cross-references the respective entries in Table 1.

Apart from the ACMG, however, most recent society guidelines pertaining to GDD/ID have recommended non-NGS technologies as first-tier genetic tests, including the following:

- Chromosomal microarray (CMA) alone (ASHG 2010 guidelines focused on DD/ID, ASD, multiple congenital anomalies ^5^)
- CMA and Fragile X testing (AAP 2014 guidelines focused on GDD/ID ^21^; AAP 2020 guidelines focused on DD ^17^)

The AAP 2020 guidelines focused on DD (including GDD and ASD) ^17^ do indicate that, for GDD/ID, “further testing [after CMA and Fragile X] may include ES and gene panels” without making a definitive recommendation.

The most recent society guidelines pertaining to ASD have also recommended non-NGS technologies as first-tier genetic tests, including:

- Karyotype, Fragile X testing, or CMA (AACAP 2014 guidelines focused on ASD ^23^)
- Karyotype (AAN and CNS 2000 guidelines focused on ASD ^10^)
- CMA and Fragile X testing (AAP 2020 guidelines focused on ASD ^20^; AAP 2020 guidelines focused on DD including GDD and ASD ^17^; ACMG 2013 guidelines focused on ASD ^22^; ASHG 2010 guidelines focused on DD/ID, ASD, and multiple congenital anomalies ^5^)

Recent guidelines on CP from the AAP 2022 ^19^ referenced genetic testing for a subset of patients (“diagnostic evaluation may include advanced genetic techniques, such as chromosomal microarray and genomic sequencing”) but did not provide detailed guidance for when this is indicated or the specific type of genetic testing that should be pursued.

## RESULTS: RECOMMENDATIONS DIVERGE ACROSS SPECIALITIES

It is notable that for a given NDD, there are differing recommendations for genetic testing depending on which society created them. These variations may arise as a result of the familiarity of providers in that specialty with new testing methodologies, consideration of the severity of the patients typically seen by that specialty, and differences in the year of publication as noted above. For example, a neurologist evaluating a patient with ASD may reference the most recent guidelines by the society pertinent to their specialty (i.e., CNS or AAN) ^10^, which would suggest karyotype and Fragile X testing as first line testing. A pediatrician seeing the same patient may reference the 2020 AAP guideline on ASD ^20^ and consider sending CMA and Fragile X testing. If the patient has co-occurring ID (a common scenario given that 30-70% of children with ASD have ID ^26^), a geneticist may adhere to the ACMG 2021 guidelines ^18^ and consider sending ES or WGS. As a result of these divergent recommendations, the same patient may undergo different sets of genetic testing depending on the referring clinicians who may be following practice guidelines most closely associated with their specialty.

## DISCUSSION: NEGATIVE IMPACT FROM LACK OF CONTEMPORARY GUIDELINES

The lack of contemporary, consistent guidelines from medical societies on genetic testing for NDDs may have a detrimental impact on patients. First, the lack of guidelines results in significant practice variability and inequitable access to care. The diagnostic yields of different testing methods for NDDs have been established through systematic reviews and meta-analyses, with ES/WGS yielding several-fold higher diagnostic rates than CMA for individuals with non-specific presentations ^6,7,12^. Utilizing a test with a lower diagnostic yield either as the sole test or as the first-line test may lead to missed or delayed diagnoses. In a 2021 national survey administered to U.S. child neurologists about genetic testing practice choices for patients with GDD/ID of unknown etiology, nearly 70% almost always pursued CMA, 60% almost always pursued Fragile X testing for male patients, 25% almost always pursued Fragile X testing for female patients, and only 11% almost always pursued ES ^27^. Although there are several possible explanations for this practice variability, the misalignment of professional society guidelines and failure to incorporate new data may contribute to practitioners choosing different first-line tests based on their familiarity with a single or small subset of available guidelines, which may be out of date. For CP, there is significant practice variability about what constitutes the very definition of CP, even among neurologists, including whether the presence of a genetic disorder *precludes* the diagnosis of CP ^13^. These studies demonstrate clear evidence that a provider’s specialty greatly impacts whether genetic testing will be done and if so, what type. Practice variability may drive inequities in care, as patients receive different evaluations based on access to specialists rather than differences in their clinical presentations.

The second potential impact from the lack of convergent, contemporary guidelines for genetic testing for NDDs is the missed opportunity for genetic diagnoses in clinical populations. ES/WGS provides a molecular diagnosis to approximately 30-40% of individuals with varying combinations of ASD, ID, and congenital anomalies ^6,28^. With each additional molecular diagnosis, there is potential benefit to the family, including termination of the diagnostic odyssey resulting in changes in medical management, surveillance, reproductive counseling, family testing, access to clinical trials and/or advocacy groups, and reduced healthcare costs ^12,28–31^. Though the reported frequency of these outcomes varies greatly due to heterogeneous study definitions, a meta-analysis found that 27%, 17%, and 6% of patients underwent a change in clinical management following results of WGS, ES, and CMA, respectively ^30^. As the number of identified NDD-related genes continues to expand with improving technology and pooling of patient samples ^32,33^, ES and especially WGS are becoming increasingly more useful than multi-gene panels. While it may seem intuitive that ES/WGS would report more variants of unknown significance (VUS) compared to multi-gene panels, the opposite has been reported, likely reflective of higher rates of concurrent parental sample availability and higher phenotypic correlation required for generating ES/WGS reports ^34^. Beyond ES/WGS, evidence supporting the clinical utility of newer technologies such as long-read WGS, transcriptomics, polygenic risk scores, and epigenetic profiling is emerging ^35–38^. Without regularly updated, comprehensive clinical genetic testing guidelines accepted by multiple specialties that care for NDD patients, patients may receive less effective, less comprehensive testing, leading to missed opportunities for genetic diagnoses and the associated benefits. The genetic landscape of NDDs is rapidly changing, and future guideline development/revision should mirror this pace. Based on this analysis and recommended previously^39^, we suggest that societies combine efforts to create a single guideline, with representation from multiple specialties, with processes in place to allow regular updates via online reports.

Inconsistent and outdated guidelines likely contribute to inequities in access to genetic care for historically-marginalized patients. Recent studies have highlighted racial discrepancies on clinical NDD diagnoses which are reasonably considered proxies for further NDD evaluation ^40–42^. Though research into disparities in access to genetic testing for NDD has been limited to date, there is evidence of inequities, with children identified as Black/African American or Hispanic less likely to receive genetic testing compared to non-Hispanic White children ^43–46^. One potential contributor to this disparity may be unequal access to specialty providers. In a retrospective study of referrals to pediatric subspecialists within one healthcare network, decreased likelihood of visit scheduling and attendance was associated with racial and socioeconomic factors, including African American race, public insurance, and lower zip code median income ^47^. With respect to genetic testing specifically, there are several factors, pertaining to both the patient and the healthcare professional, that impede a referral to genetic services, ^48^. Thus, society guidelines failing to promote the highest yield test as the first-tier test may lead to additional visits as part of the diagnostic odyssey, potentially widening existing health disparities for individuals with NDDs ^49,50^.

Insurance coverage for genetic testing in NDDs also is critically hindered by outdated clinical guidelines, resulting in underutilization of evidence-based diagnostically proven tools such as ES. Clinicians often recognize best practices but run into limitations in implementation. For example, 65% of U.S. child neurologists support ES as a first-line diagnostic test, but only about 10% routinely obtain it, often due to insurance constraints ^27^. Furthermore, in a sample of 4,500 prior authorizations for genetic testing in Texas, one-third of ES requests were denied due to lack of medical necessity, characterization of such testing as experimental/investigational, or not meeting criteria for testing per the payer rules ^51^. Interestingly, in the Texas study, the inclusion of specific diagnostic codes when submitting a prior authorization request did not influence the outcome of the prior authorization decision, *except* in the instance of the International Classification of Diseases, Tenth Revision (ICD-10) code for Autism (F84.0) in requests for CMA testing, which was associated with a higher approval rate ^51^. This potentially reflected payers’ incorporation of outdated society guidelines recommending CMA as a first-line test for ASD within their own coverage plans.

There is limited research into how payers develop coverage rules for pediatric ES/WGS. One qualitative study conducted in 2019 found that 70% of payers provided coverage for pediatric ES, though most felt there was “insufficient evidence of clinical utility” ^52^. Presumably, consistent and updated clinical societal guidelines would support endorsement of the clinical utility of genetic testing and may lead to increased coverage by payers. Within oncology genetic testing and payer coverage, there has been a call for incorporating regularly-updated society guidelines in the health technology assessment process used in coverage decision-making ^53^. A similar approach should be considered for NDDs.

## RECOMMENDATIONS

Based on the above results, examples from adjacent clinical fields, and the authors’ collective expertise in the care of children with NDDs, we have created a series of recommendations for medical societies engaged in the development or updating of genetic testing practice guidelines.

1. **Professional societies should combine efforts to create a single guideline.** Such an approach can be accomplished by including members from multiple specialties, potentially providing more uniformity between practitioners caring for individuals with NDD. Furthermore, such a process can be updated via online reports reducing redundancy intrinsic to the current guideline development processes of different societies.
2. **Medical societies should formally endorse other societies’ genetic testing practice guidelines.** Endorsement of an existing guideline may minimize duplication of effort while promoting cross- or intra-disciplinary collaboration. For example, with respect to epilepsy, the National Society of Genetic Counselors (NSGC) published a practice guideline in 2023 recommending ES/WGS and/or a multi-gene panel (>25 genes) as a first-tier test, followed by CMA if the initial testing was non-diagnostic ^54^. The American Epilepsy Society has also endorsed this guideline. This endorsement increases the guideline’s impact by promoting awareness among members of both societies. Endorsement of existing guidelines for genetic testing for an NDD does not preclude the creation or updating of separate guidelines that focus on other aspects of the NDD, like diagnosis and management.
3. **Medical societies should consider creating/updating practice guidelines for genetic testing separately from guidelines that are disorder-specific and focused on other aspects of diagnosis or management.** For example, the ACMG 2021 guidelines recommend ES/WGS as a first- or second-tier test for all individuals with DD/ID and/or congenital anomalies ^18^. These guidelines outline the evidence behind this genetic testing recommendation but do not discuss other aspects of care, such as screening, imaging, other lab work, medications, or surveillance for associated conditions. In contrast, the most recent AAP guidelines on developmental surveillance and screening embed a discussion of genetic testing for children with DD ^17^. A separate statement on genetic testing specifically would allow for easier, more frequent updates and provide a quicker reference point to clinicians.
4. **Medical societies should strongly consider the genetic heterogeneity and phenotypic overlap of NDDs in the development of genetic testing guidelines.** The number of genes associated with NDDs continues to rapidly expand, as do the phenotypes of individual genetic disorders (e.g., MECP2-related disorders, GLUT1 deficiency, among other examples). Due to this, we suggest broad spectrum evaluation with ES/WGS as first-line testing for individuals with unexplained NDDs and non-specific features.
5. **Medical societies should structure their guidelines to inform payer coverage policies that are aligned with clinical best practices.** Given that payer medical policies often cite clinical guidelines ^55^, medical societies should be aware of how their guidelines influence access to testing. Guidelines should clearly delineate the clinical benefits of genetic testing and the ability of such testing to guide clinician decision-making and recommendations for follow-up care, as well as the value of genetic testing to patients and families. Given the quick pace of progress in genetic technologies, payers would benefit from additional clinical input, particularly regarding the diagnostic sensitivity of various assays for genomic anomalies of interest ^56^. We recommend medical societies notify payers when both systematic evidence reviews and evidence-based practice guidelines are published in an effort to promote timely development of evidence-based payer policies and reduce coverage-based disparities in access to genetic testing.
6. **Genetic testing practice guidelines should specifically address recommendations for improving access among marginalized patient populations.** Further research is required to investigate reasons for disparities in access to and uptake of genetic testing among marginalized population groups and the relationship between access to genetic testing and structural determinants of health. However, societies should acknowledge potential barriers that disproportionately affect marginalized groups, and as more research accumulates, suggest targets for intervention to increase equitable access.
7. **Medical societies with guidelines pertaining to genetic testing should strive to update or reaffirm guidelines every 2 years.** This cadence, while frequent, is reflective of the rapid evolution of genetic/genomic technologies. If a society is unable to reaffirm or update a specific guideline, it can endorse more updated guidelines from another society in the interim. The importance of frequent updates to genetics-related practice guidelines is underscored by the rapid pace of genomic discoveries and technologies over the past two decades. Dozens of genes are discovered each year to be associated with disorders in the context of pathogenic variants ^57^. On a daily basis, approximately 10 new genetic tests are made available in the U.S. and global markets ^58,59^.
8. **An online resource should be created to track ongoing recommendations for genetic testing for NDDs.** This could be modeled off the National Comprehensive Cancer Network (NCCN)’s extensive online guideline process, which includes guidelines related to genetic testing and involves continual review and updates to ensure they are reflective of current evidence ^60^. This approach would ensure that NDD society guidelines include the most updated evidence and are quickly accessible to clinicians and payers. Ideally, these guidelines would be reflected in coverage plans across all insurance providers, which could minimize the contribution of insurance status to inequities in access to genetic testing.

## LIMITATIONS

Our work has a few notable limitations. First, we focused exclusively on U.S.-based genetic testing guidelines due to the differences between healthcare systems and payment structures in other countries. Nonetheless, the principles outlined in this review, including the need for modernized guidelines across specialties for different NDDs, may still be relevant to healthcare systems in different countries. Second, we limited our scoping review to include only GDD/ID, ASD, and CP, three major NDDs for which there is scientific evidence regarding the utility of genetic testing. Over time, our scientific knowledge may expand to include an understanding of the yield of genetic testing for other NDDs, such as ADHD^61^, reiterating the need to continually update and revise society guidelines. We did not evaluate primary data used to develop these guidelines.

## CONCLUSION

Genetic testing remains a rapidly evolving field with falling costs and improved diagnostic yields. Like other clinical guidelines, an appropriate balance must be struck between the current evidence base and flexibility to remain relevant over time. As the applications of genetic testing have greater impact on early diagnosis and intervention, there must be a greater involvement of all stakeholders, including payers, policymakers, and family advocacy groups.

## Data Availability

Data from this scoping review is available upon request from the corresponding author.

## ADDITIONAL INFORMATION

## Acknowledgements

We would like to acknowledge Dr. Tracy King at NICHD for her facilitation of the IDDRC Workgroup on Advocating for Access to Genomic Testing.

This study was made possible by the following IDDRC grants: P50HD105351 (Boston Children’s Hospital), P50HD103538 (Kennedy Krieger Institute), P50HD103556 (University of Iowa), P50HD103537 (Vanderbilt University), and P50HD103525 (Washington University in St. Louis).

SS receives support from the National Institute of Health/National Institute of Neurological Disorders and Stroke (NIH/NINDS) (1K23NS119666). JJC receives research support from the NIH/NINDS (5K12NS098482). JSC receives funding from the National Institute of Child Health and Human Development (NICHD) (P50 HD103538), NINDS (1U24NS131172-01), and U.S. Department of Health and Human Services, Health Resources and Services Administration (HRSA) (MCH T7317245). MAD receives funding from the NICHD (R21HD101977), National Human Genome Research Institute (NHGRI) (R21HG012626) and HRSA (HRSA-22-134). HSS receives funding from the NHGRI (R00HG011491). EP receives support from the Cincinnati Children’s Hospital Medical Center Research Foundation, NIH National Institute of Mental Health (NIMH) (R01-HD108222, U01-DA055342), Center for Disease Prevention (CDC) (U01-CE003570), NICHD (U54-HD104461), and NIH/NINDS (R01-NS117597). JSA receives funding from Autism Speaks (Autism Care Network). MS receives funding from the NIH (P50HD105351, U54NS092090). CAG receives support from the National Institute of Arthritis and Musculoskeletal and Skin Diseases (NIAMS) (R01AR067715), NICHD (P50HD103525), and National Center for Advancing Translational Sciences (NCATS) (UL1TR000448). No funding organization had a role in the study design, implementation, or analysis.

## Author Contributions

All authors contributed to the conception and design of the study. SS and JJC performed the acquisition and analysis of the data. SS, JJC, and JSC drafted a significant portion of the manuscript or figures. All authors contributed significant intellectual content to the study and provided critical edits to the manuscript. Please see Supplementary Table 1 for a complete list of members of the IDDRC Workgroup on Advocating for Access to Genomic Testing.

## Data Availability

Data from this scoping review is available upon request from the corresponding author.

