## Supplementary material for "Survey of the Landscape of Society Practice Guidelines for Genetic Testing of Neurodevelopmental Disorders": Table S1

Supplementary Table 1: Membership of the IDDRC Workgroup on Advocating for Access to Genomic Testing

| **Name, Degree(s)** | **Specialty** | **Affiliation** |
| --- | --- | --- |
| Julia Anixt, MD | Developmental Behavioral Pediatrics | Department of Pediatrics, University of Cincinnati College of Medicine; Cincinnati Children’s Hospital |
| Kayla Blankenship, MS, LGC | Genetics | Department of Neurology, University of Iowa |
| Tashalee Brown, MD, PhD | Child Psychiatry | Department of Psychiatry, University of California Los Angeles |
| Colleen Campbell, PhD, MS, LGC | Genetics | Department of Internal Medicine, University of Iowa |
| Maya Chopra, MBBS, FRACP | Genetics | Department of Neurology, Boston Children’s Hospital |
| Julie S. Cohen, ScM, CGC | Genetics | Department of Neurology and Developmental Medicine, Kennedy Krieger Institute; Department of Neurology, Johns Hopkins University School of Medicine |
| Jordan J. Cole, MD | Child Neurology | Department of Pediatrics, University of Colorado |
| Brian Corner, MS, CGC | Genetics | Department of Pediatrics, Vanderbilt University |
| Matthew Deardorff, MD, PhD | Genetics | Departments of Pathology and Laboratory Medicine & Pediatrics, University of Southern California |
| Tiffany Grider, MS, LGC | Genetics | Department of Neurology, University of Iowa |
| Christina Gurnett, MD, PhD | Child Neurology | Department of Neurology, Washington University in St. Louis |
| Elizabeth Jalazo, MD | Genetics | Department of Pediatrics, University of North Carolina |
| Shafali Jeste, MD | Child Neurology | Departments of Neurology & Pediatrics, University of Southern California |
| Kate MacDuffie, PhD, MA | Psychology/Bioethics | Department of Pediatrics, University of Washington |
| Ernest Pedapati, MD, MS | Child Psychiatry | Department of Psychiatry and Behavioral Neuroscience, Cincinnati Children’s Hospital |
| Mustafa Sahin, MD, PhD | Child Neurology | Department of Neurology, Boston Children’s Hospital |
| Angela Sellitto, MS | Clinical Research Coordinator | Department of Neurology, Washington University in St. Louis |
| Hadley Smith, PhD, MPSA | Health economics | Department of Population Medicine, Harvard Medical School and Harvard Pilgrim Health Care Institute |
| Siddharth Srivastava, MD | Child Neurology | Department of Neurology, Boston Children’s Hospital |
| Olivia Veatch, PhD | Biomedical informatics | Department of Psychiatry and Behavioral Sciences, University of Kansas |
